# Intraepithelial CD15 infiltration identifies high grade anal dysplasia in people with HIV

**DOI:** 10.1101/2023.08.17.23294199

**Authors:** Joaquín Burgos, Cristina Mancebo, Núria Massana, Antonio Astorga-Gamaza, Josep Castellvi, Stefania Landolfi, Adrià Curran, Jorge N. Garcia-Perez, Vicenç Falcó, María J. Buzón, Meritxell Genescà

**Author notes:** Correspondence (J.B.) and (M.G.). These authors contributed equally (Cristina Mancebo, Núria Massana). The authors declare no potential conflicts of interest.

## Abstract

Men who have sex with men (MSM) with HIV are at high risk for squamous intraepithelial lesion (SIL) and anal cancer. The identification of local immunological mechanisms involved in the development of anal dysplasia could aid treatment and diagnostics. We performed a study of 111 anal biopsies obtained from 101 MSM with HIV, who participated in an anal screening program. In a test prospective cohort (N=54), in addition to histological examination, we assessed multiple immune subsets by flow cytometry. Selected molecules were further evaluated by immunohistochemistry in a validation retrospective cohort (N=47). Pathological samples were characterized by the presence of Resident Memory T cells with low expression of CD103 and by changes in the Natural Killer cell subsets, affecting residency and activation. Furthermore, potentially immune suppressive subsets, including CD15^+^ CD16^+^ mature neutrophils, gradually increased as the anal lesion progressed. Immunohistochemistry confirmed the association between the presence of CD15 in the epithelium and SIL diagnosis, with a sensitivity of 80% and specificity of 71% (AUC 0.762) for the correlation with high-grade SIL. A complex immunological environment with imbalanced proportions of resident effectors and immune suppressive subsets characterizes pathological samples. Neutrophil infiltration, determined by CD15 staining, may represent a valuable pathological marker associated with the grade of dysplasia.

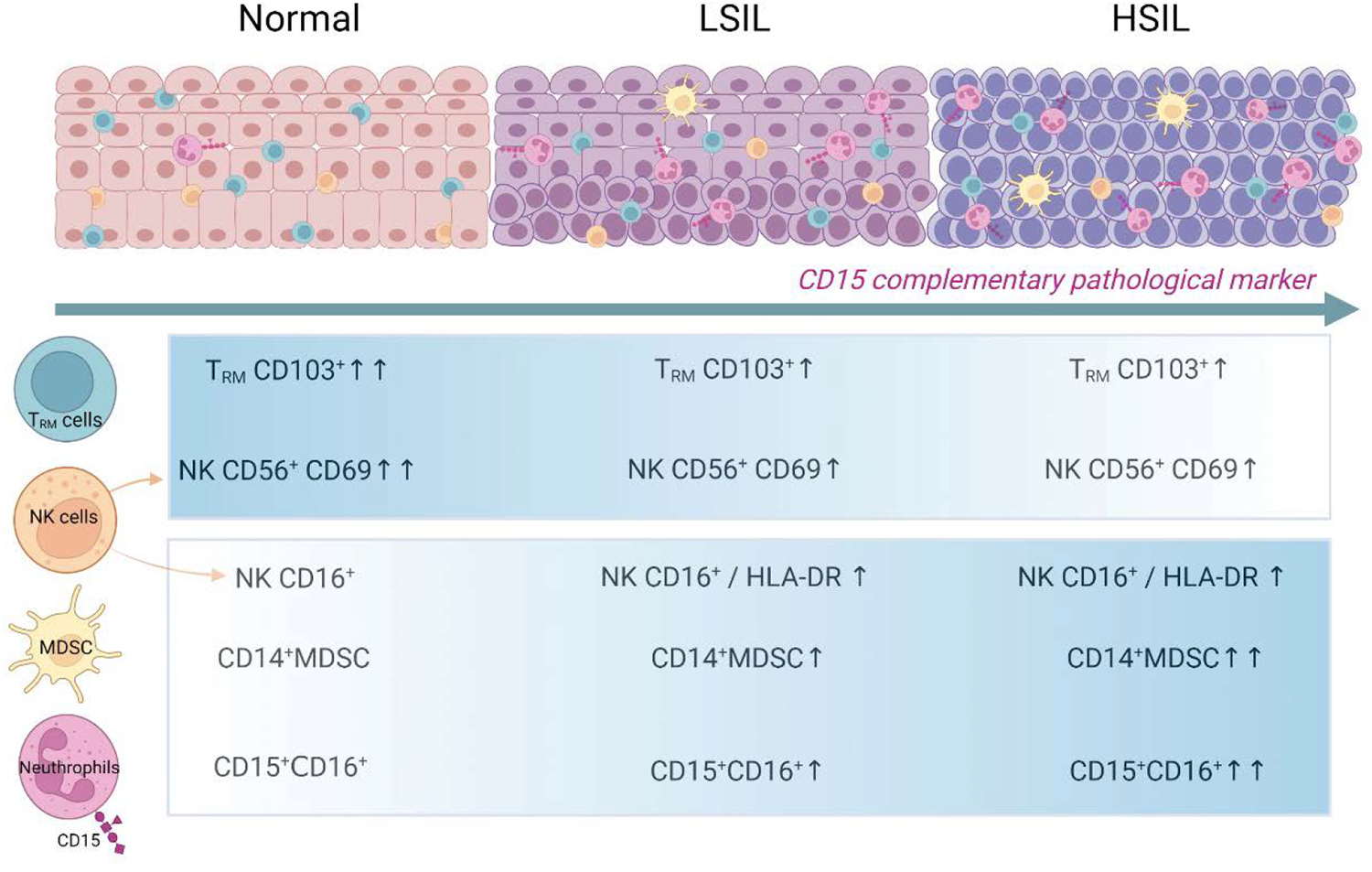

## INTRODUCTION

Anal cancer is considered infrequent in the general population (1). However, in selected populations, as men who have sex with men (MSM) with HIV, anal cancer occurs at rising rates and is currently one of the most common non-AIDS defining cancers (2). Infection by high-risk human papillomavirus (HR-HPV) at the squamocolumnar transition zone (TZ) is considered the main etiological agent of anal cancer (3). Persistent HPV infection is able to induce a series of changes in the transitional epithelium that lead to the development of low-grade squamous intraepithelial lesion (LSIL), which in turn progresses to high-grade squamous intraepithelial lesion (HSIL), considered to be the direct precursor of invasive anal cancer (4, 5).

Anal squamous intraepithelial lesions (SIL) are histologically identical among people with HIV (PWH) and uninfected individuals; however, they are more prevalent and likely to persist and progress to anal cancer in the first group, even among those in which antiretroviral therapy (ART) maintains viral suppression and induces immunological recovery (2, 6). Multiple factors related to the local interaction and potentiation between HIV and HPV may explain this increase in prevalence and associated-pathology in PWH, including oncogenic effects and overall impact on local immunity (7–10). Of particular importance may be the persistent depletion of CD4^+^ T cells from the mucosal compartments in PWH who have been treated during chronic infection (11, 12), which may create a more favorable microenvironment for precancerous lesions to develop and progress. In this sense, altered cell-mediated immunity has been associated with increased HPV infection and disease (13), while immune responses orchestrate regression of HPV-related lesions (14).

Screening and treating HSIL have recently been demonstrated to be effective for cancer prevention in PWH (15). However, a reliable biomarker that indicates the risk to develop anal cancer has not yet been identified, and even classifying intermediate lesions of SIL is still challenging (16). Most studies attempting to expand the understanding of anal dysplasia progression and find potential biomarkers have focused on the genes and/or proteins involved in HPV-mediated carcinogenesis (17, 18). In contrast, studies focusing on the local immune microenvironment surrounding anal lesions are scarce (19), although disturbances in the local microenvironment may play a critical role in the development of anal cancer precursors (20, 21). Thus, phenotyping the immune landscape surrounding dysplastic lesions could provide new insights on the immunopathology of these persistent infections, which in turn may allow the identification of new biomarkers.

In light of the limited data available about the immune microenvironment that differentiate normal epithelium from anal dysplastic lesions and the limitations of diagnostic tools of HSIL, we conducted a study to evaluate immunological subsets in the anal mucosa of MSM with HIV who participated in an anal screening program. The main goal of this study was to characterize the immune environment where lesions develop to identify biomarkers that can contribute to diagnose HSIL.

## METHODS

### Study design and patient cohorts

The Anal Dysplasia Unit at the University Hospital Vall d’Hebron (HUVH, Barcelona, Spain) was created in May 2009 and attends more than 1,000 MSM with HIV. Anal screening program includes anal liquid cytology and HPV determination, a high-resolution anoscopy (HRA) and, when necessary, anal biopsies, as previously described (22). Patients undergoing anal biopsies as part of the screening program were offered to participate in the study with the following inclusion criteria: patients on ART, with HIV viral suppression and without any anal sexually transmitted diseases in the last 6 months. Patients were included prospectively for the initial immunological and histological analyses, while for the validation of the results by immunohistochemistry, patients were recruited retrospectively from available histological samples. Informed consent for sample collection and use of information available in the medical records was obtained from all patients included. This study was performed in accordance with the Declaration of Helsinki and approved by the Institutional Review Board (PR(AG)240/2014) of the HUVH.

### Sample collection

Cytology was obtained by introducing a Dacron swab 3-5cm into the anal canal and softly rotating it. The swab was introduced into 20ml of PreservCyt/ThinPrep Pap test solution (Cytyc Iberia SL, Barcelona, Spain) and shaken for 30 seconds. This sample was used to carry out the cytological analysis and HPV testing (22). Single or multiple anal biopsies were taken from individual patients in the same screening session if HRA revealed an abnormal area and/or in areas that were previously treated to determine treatment efficacy. For a single biopsy, an immunological and histological study was carried out simultaneously. An expert pathologist classified samples using the terminology and morphological criteria published in the Lower Anogenital Squamous Terminology (LAST) project: benign, LSIL and HSIL (16, 23).

### HPV detection

DNA was extracted from cytology-derived cell suspensions using the “QIAamp Viral DNA minikit” (QIAGEN, Hilden, Germany). Specific sequences of papillomavirus were amplified by specific protocol “CLART® Genomic HPV-2” in accordance with the manufacturer’s instructions. Detection of HPV genotypes 16, 18, 31, 33, 35, 39, 45, 51, 52, 56, 58, 59, 68, 73, and 82 were considered as high-risk.

### Immunological cell phenotyping by cytometry

Fresh anal tissue samples of ≈8mm^3^ were collected in antibiotics-containing RPMI 1640 medium. Samples were enzymatically digested with 5mg/ml collagenase IV (Gibco) and the resulting mononuclear cells suspension was washed twice and stained for viability with Live/Dead Aqua (Invitrogen) at room temperature for 30min in phosphate-buffered saline (PBS). Cells were then washed with PBS and surface stained using a 13-color flow cytometry panel (**Supplementary Table 1**). After fixation, all events were acquired using a BD LSRFortessa flow cytometer and data was analysed with FlowJo vX.0.7 software (TreeStar). We established a minimum of 1% of CD45^+^ cells from the total stored events as well as additional minimums per subset count to consider the sample for the analyses: 100 events for CD3^+^T-cell lymphocytes, 50 events for CD3^-^ lymphocytes and 100 events for myeloid cells.

### Immunohistochemistry

We analyzed CD103- and CD15-positive cells by immunohistochemistry to assess their value as a pathology marker in archival specimens from anal biopsies obtained from the validation cohort. Formalin-fixed, paraffin-embedded anal samples of 3-μm sections were deparaffined, rehydrated and were stained using optimal dilutions of monoclonal antibodies (**Supplementary Table 1**). The staining was performed following the protocol of the *ultraView Universal DAB* kit for Ventana Benchmark ultra. Mononuclear cells with a dark brown cytoplasmic signal were recorded as positive cells. Since intensity of the staining was homogeneous, the H score (an indicator of the intensity and proportion of the biomarker identified) was not used. Positive cells within the squamous epithelium and underlying stroma of the whole sample were manually counted using light microscopy at 40× magnification by two independent pathologists (J.C and S. L). Sections were examined avoiding lymphoid follicles of the stromal areas when present. The average number of positive cells from a median of 3 fields was reported for all the markers except for p16, which was considered positive or negative based on the staining at the nuclear level of the squamous epithelial cells.

### Statistical Analysis

Comparisons were performed between the three histological groups (normal, LSIL and HSIL) as well as between two groups (normal *versus* pathological samples, which combined LSIL and HSIL samples) to increase statistical power. Statistical analysis was conducted using GraphPad Prism software. Non-parametric Kruskal-Wallis test with Dunn’s post-hoc test for multiple comparisons and Mann-Whitney U test or Chi-square test were used for the unpaired analyses of three and two groups, respectively. For patients with paired normal and a LSIL samples, we employed the Wilcoxon signed-rank test. Sensitivity, specificity, and the area under the receiver operating characteristic (ROC) curve of potential biomarkers to detect HSIL were also determined in the validation cohort.

## RESULTS

### Cohort characteristics

The prospective cohort included 47 ART-treated, MSM with HIV with a total of 54 histological anal samples in which flow cytometry analyses were performed. Anal samples were histological classified as normal (n=24 samples, including 19 individuals), LSIL (n=24 samples, including 19 individuals) and HSIL (n=6 samples, including 6 individuals). In 7 of these individuals we had concomitant paired samples, wherein one was classified as normal and the other as LSIL. The validation cohort included 54 MSM with HIV with 57 anal samples, which were classified as normal (n=12), LSIL (n=25) and HSIL (n=20). **Table 1** shows a summary of the participant characteristics related to HIV and other relevant parameters from both cohorts.

**Table 1.**
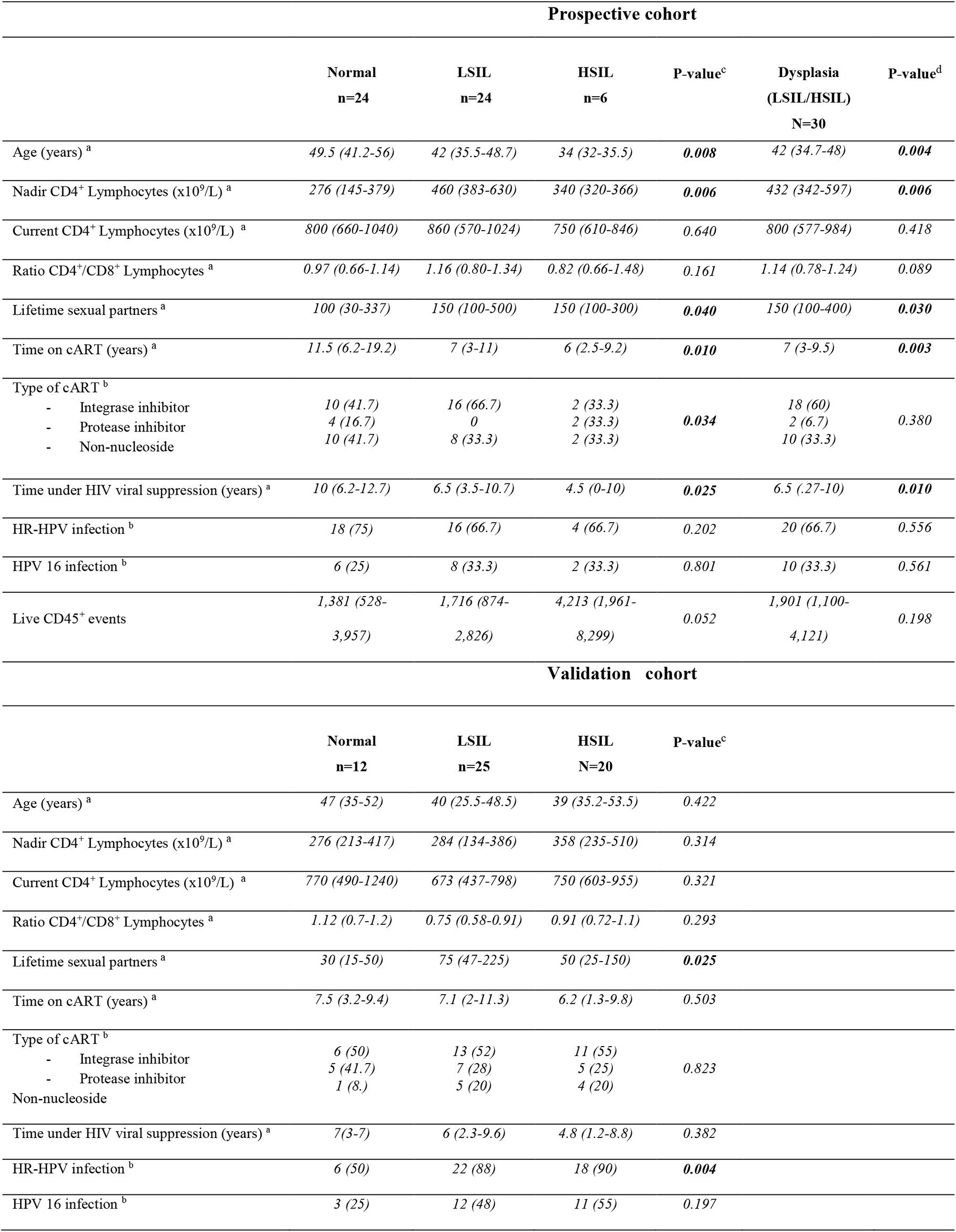
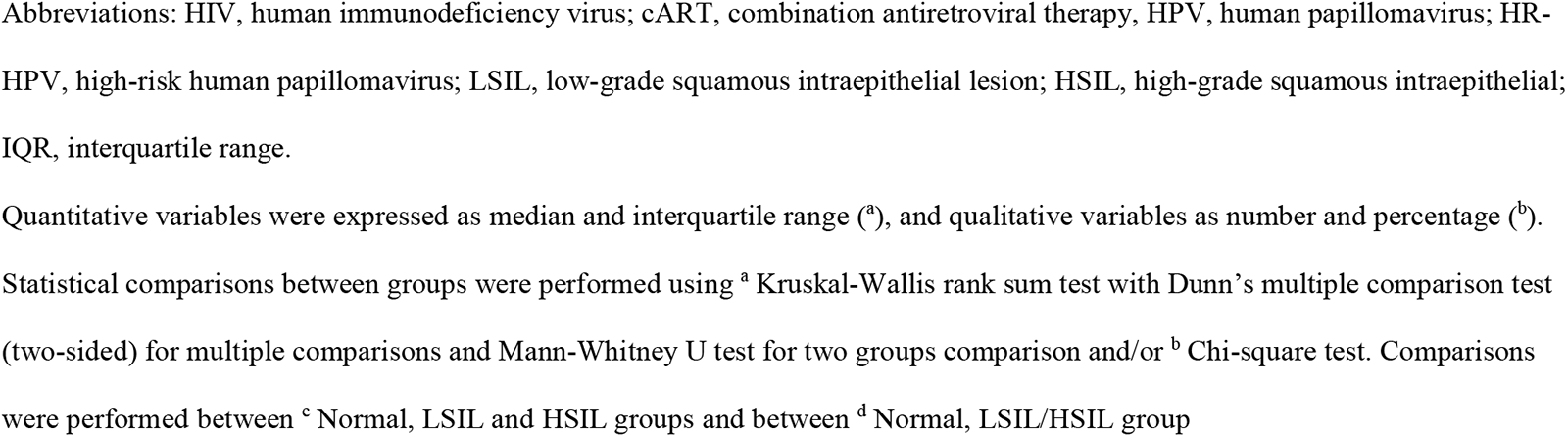
Patient characteristics.

| Prospective cohort |  |  |  |  |  |  |
| --- | --- | --- | --- | --- | --- | --- |
|  | Normal<br>n=24 | LSIL<br>n=24 | HSIL<br>n=6 | P-value<sup>c</sup> | Dysplasia<br>(LSIL/HSIL)<br>N=30 | P-value<sup>d</sup> |
| Age (years) <sup>a</sup> | 49.5 (41.2-56) | 42 (35.5-48.7) | 34 (32-35.5) | **0.008** | 42 (34.7-48) | **0.004** |
| Nadir CD4<sup>+</sup> Lymphocytes (x10<sup>9</sup>/L) <sup>a</sup> | 276 (145-379) | 460 (383-630) | 340 (320-366) | **0.006** | 432 (342-597) | **0.006** |
| Current CD4<sup>+</sup> Lymphocytes (x10<sup>9</sup>/L) <sup>a</sup> | 800 (660-1040) | 860 (570-1024) | 750 (610-846) | 0.640 | 800 (577-984) | 0.418 |
| Ratio CD4<sup>+</sup>/CD8<sup>+</sup> Lymphocytes <sup>a</sup> | 0.97 (0.66-1.14) | 1.16 (0.80-1.34) | 0.82 (0.66-1.48) | 0.161 | 1.14 (0.78-1.24) | 0.089 |
| Lifetime sexual partners <sup>a</sup> | 100 (30-337) | 150 (100-500) | 150 (100-300) | **0.040** | 150 (100-400) | **0.030** |
| Time on cART (years) <sup>a</sup> | 11.5 (6.2-19.2) | 7 (3-11) | 6 (2.5-9.2) | **0.010** | 7 (3-9.5) | **0.003** |
| Type of cART <sup>b</sup> |  |  |  |  |  |  |
| - Integrase inhibitor | 10 (41.7) | 16 (66.7) | 2 (33.3) | **0.034** | 18 (60) | 0.380 |
| - Protease inhibitor | 4 (16.7) | 0 | 2 (33.3) |  | 2 (6.7) |  |
| - Non-nucleoside | 10 (41.7) | 8 (33.3) | 2 (33.3) |  | 10 (33.3) |  |
| Time under HIV viral suppression (years) <sup>a</sup> | 10 (6.2-12.7) | 6.5 (3.5-10.7) | 4.5 (0-10) | **0.025** | 6.5 (.27-10) | **0.010** |
| HR-HPV infection <sup>b</sup> | 18 (75) | 16 (66.7) | 4 (66.7) | 0.202 | 20 (66.7) | 0.556 |
| HPV 16 infection <sup>b</sup> | 6 (25) | 8 (33.3) | 2 (33.3) | 0.801 | 10 (33.3) | 0.561 |
| Live CD45<sup>+</sup> events | 1,381 (528-3,957) | 1,716 (874-2,826) | 4,213 (1,961-8,299) | 0.052 | 1,901 (1,100-4,121) | 0.198 |
| Validation cohort |  |  |  |  |  |  |
|  | Normal<br>n=12 | LSIL<br>n=25 | HSIL<br>N=20 | P-value<sup>c</sup> |  |  |
| Age (years) <sup>a</sup> | 47 (35-52) | 40 (25.5-48.5) | 39 (35.2-53.5) | 0.422 |  |  |
| Nadir CD4<sup>+</sup> Lymphocytes (x10<sup>9</sup>/L) <sup>a</sup> | 276 (213-417) | 284 (134-386) | 358 (235-510) | 0.314 |  |  |
| Current CD4<sup>+</sup> Lymphocytes (x10<sup>9</sup>/L) <sup>a</sup> | 770 (490-1240) | 673 (437-798) | 750 (603-955) | 0.321 |  |  |
| Ratio CD4<sup>+</sup>/CD8<sup>+</sup> Lymphocytes <sup>a</sup> | 1.12 (0.7-1.2) | 0.75 (0.58-0.91) | 0.91 (0.72-1.1) | 0.293 |  |  |
| Lifetime sexual partners <sup>a</sup> | 30 (15-50) | 75 (47-225) | 50 (25-150) | **0.025** |  |  |
| Time on cART (years) <sup>a</sup> | 7.5 (3.2-9.4) | 7.1 (2-11.3) | 6.2 (1.3-9.8) | 0.503 |  |  |
| Type of cART <sup>b</sup> |  |  |  |  |  |  |
| - Integrase inhibitor | 6 (50) | 13 (52) | 11 (55) | 0.823 |  |  |
| - Protease inhibitor | 5 (41.7) | 7 (28) | 5 (25) |  |  |  |
| - Non-nucleoside | 1 (8.) | 5 (20) | 4 (20) |  |  |  |
| Time under HIV viral suppression (years) <sup>a</sup> | 7(3-7) | 6 (2.3-9.6) | 4.8 (1.2-8.8) | 0.382 |  |  |
| HR-HPV infection <sup>b</sup> | 6 (50) | 22 (88) | 18 (90) | **0.004** |  |  |
| HPV 16 infection <sup>b</sup> | 3 (25) | 12 (48) | 11 (55) | 0.197 |  |  |

Abbreviations: HIV, human immunodeficiency virus; cART, combination antiretroviral therapy, HPV, human papillomavirus; HR-HPV, high-risk human papillomavirus; LSIL, low-grade squamous intraepithelial lesion; HSIL, high-grade squamous intraepithelial; IQR, interquartile range.
Quantitative variables were expressed as median and interquartile range (<sup>a</sup>), and qualitative variables as number and percentage (<sup>b</sup>).
Statistical comparisons between groups were performed using <sup>a</sup> Kruskal-Wallis rank sum test with Dunn's multiple comparison test (two-sided) for multiple comparisons and Mann-Whitney U test for two groups comparison and/or <sup>b</sup> Chi-square test. Comparisons were performed between <sup>c</sup> Normal, LSIL and HSIL groups and between <sup>d</sup> Normal, LSIL/HSIL group

### Expression of CD103 in resident memory lymphocytes is diminished in pathological samples

In order to study immune populations located in the anal biopsies, after selecting live single CD45^+^ cells, we delineated three major subsets: T lymphocytes, NK cells and specific myeloid populations. The flow cytometry gating strategy used for all samples is shown in **Supplementary** Figure 1. The median number of live hematopoietic CD45^+^cells recovered from each biopsy sample is shown in **Table 1**. Regarding the analyses of T cells derived from anal biopsies, **Figure 1A** displays two representative samples showing the frequency of CD4^+^ (here defined as CD8^-^ CD3^+^ T cells) and CD8^+^ CD3^+^ T cell subsets analyzed in normal and HSIL biopsies. In these subsets, we determined lymphocyte activation by HLA-DR expression and tissue residency by CD69 combined with CD103 expression (11). Although the overall frequency of CD4 or CD8 T lymphocytes did not vary significantly among the different groups, analyzing the total frequency of CD8^+^ T cells in paired samples from the same individual, in cases where both normal and LSIL biopsies were available, revealed a higher total frequency in LSIL samples compared to normal samples (p = 0.031; **Figure 1B**). In contrast, the fraction of CD8^+^ resident memory T cells (T_RM_) expressing CD103^+^ decreased with an increase in the degree of pathology, showing a trend when comparing normal and HSIL samples (p = 0.08, Figure 1C). Indeed, when considering pathological samples as a single group, the trend for CD8^+^ T_RM_ CD103^+^ cells remained (p = 0.063 **Figure 1D**), while CD4^+^ T_RM_ CD103^+^ cells were significantly lower in pathological samples compared to non-pathological biopsies (p = 0.024; **Figure 1 E**). Thus, while CD8^+^ T cell infiltration appeared to be associated with a LSIL diagnostic, the proportion of T_RM_ expressing CD103 was reduced as the level of dysplasia progressed. This reduction was more pronounced for CD4^+^ T_RM_ lymphocytes.

**Figure 1.**
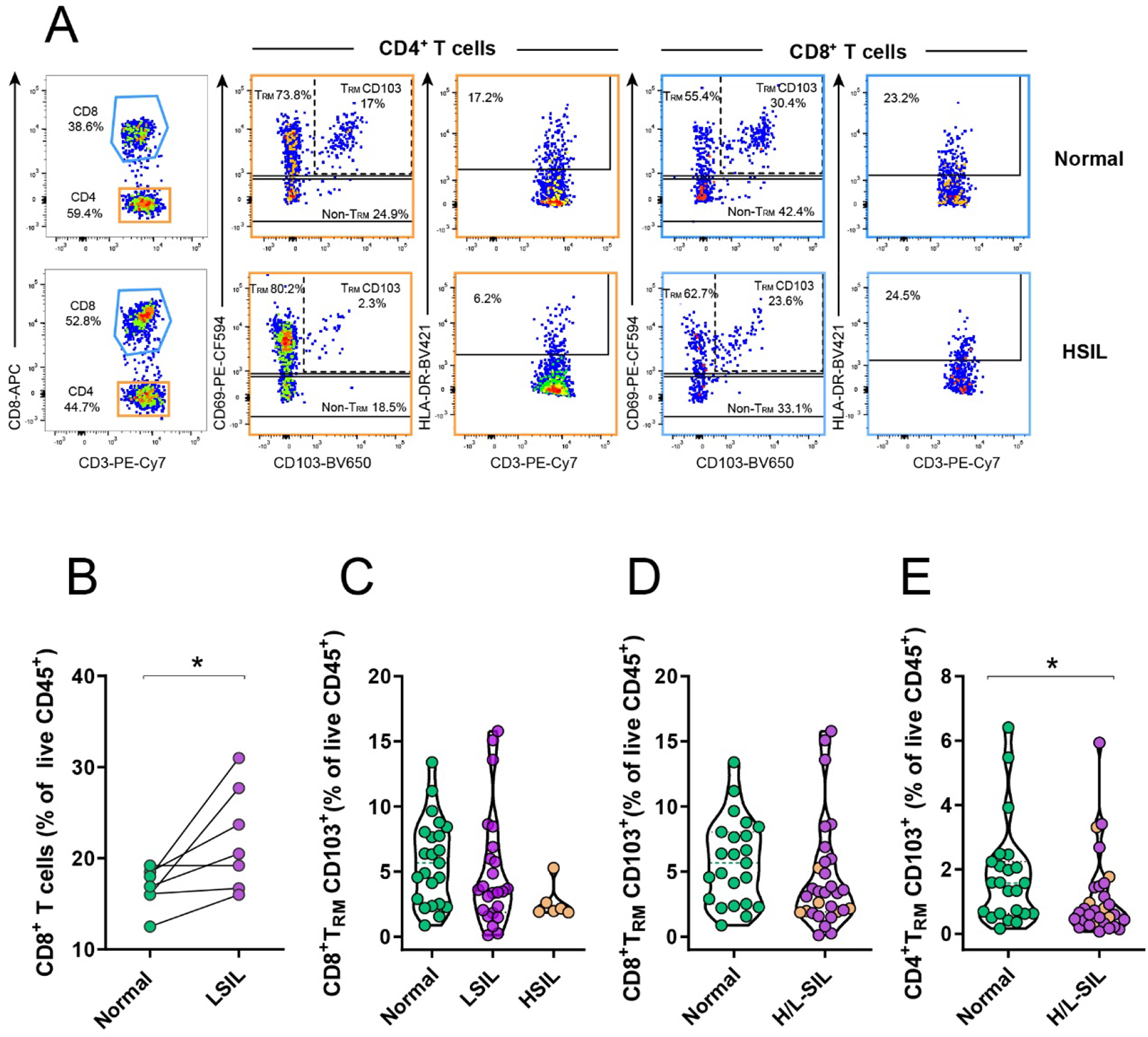
Changes in CD3^+^T lymphocyte populations associated to squamous intraepithelial lesion. **A**, Examples of the gating strategies used to identify T cell subsets in a normal anal sample (top) and in a high grade squamous intraepithelial lesion sample (HSIL, bottom). Sequential gating from left to right was used to identify four populations of interest within CD4^+^ T cells (in orange) or CD8^+^ T cells (in blue): T_RM_ (CD69^+^CD103^-^) T_RM_CD103^+^, non-T_RM_ (CD69^-^CD103^-^) and activated HLA-DR^+^ T cells. **B,** Frequencies of total CD8^+^T lymphocytes in paired concomitant normal and LSIL samples from the same individual analysed by the Wilcoxon signed-rank test. *p<0.05. **C and D**, Frequencies of CD8^+^ T_RM_ CD103^+^ lymphocytes out of all living CD45^+^ cells in (**B**) the three study groups or in (**C**) normal (in green) *versus* pathological (H/L-SIL, in purple; HSIL are highlighted in brown) samples. **E**, Frequencies of CD4^+^T_RM_ in normal (in green) *versus* pathological (H/L-SIL, in purple; HSIL are highlighted in brown) samples. Data are represented as a violin plot; horizontal lines are median and interquartile range. Statistical comparisons using non-parametric Kruskal-Wallis test with Dunn’s post-hoc test for multiple comparisons and Mann-Whitney U test for two group analyses are shown: *p<0.05.

### Natural Killer cells expressing CD56 are perturbed in pathological samples

We then analyzed the frequency CD3^-^ lymphocytes based on their expression of CD16 or CD56, as the major NK subsets in tissue samples, as shown in the representative examples (**Figure 2A**). Of note, since biopsies were small and the number of cells analyzed highly limited (in particular within the CD3^-^ lymphocyte subset, in which we considered a minimum of 50 cells), we did not attempt to analyze all the potential combinations of dim vs high CD16/CD56 NK subsets. Overall, a larger percentage of CD16^+^ NK cells tended to accumulate in the LSIL biopsies compared to normal samples (p = 0.078); an observation that was significant when we compared all pathological samples with the non-pathological ones (p = 0.030, **Figure 2B**). For each CD56 or CD16 NK cell subset, we also analyzed the expression of markers associated with residency in tissue (CD69) (24), or with cellular activation (HLA-DR) (**Figure 2A**). Interestingly, both pathological and normal samples showed very high expression of CD69 within the CD56^+^ NK subset, and this expression was higher in the non-pathological group compared to the pathological one (p = 0.041, **Figure 2C**). In contrast, in terms of HLA-DR expression, pathological biopsies showed significantly higher percentages of this molecule in the CD56^+^ NK cell fraction compared to normal biopsies (p = 0.031, **Figures 2D**). These results indicate that CD16^+^ NK cells and HLA-DR expression within the CD56^+^ NK subset is augmented during pathology, while the frequency of CD56^+^ NK cells expressing CD69 may be compromised in dysplastic environments.

**Figure 2.**
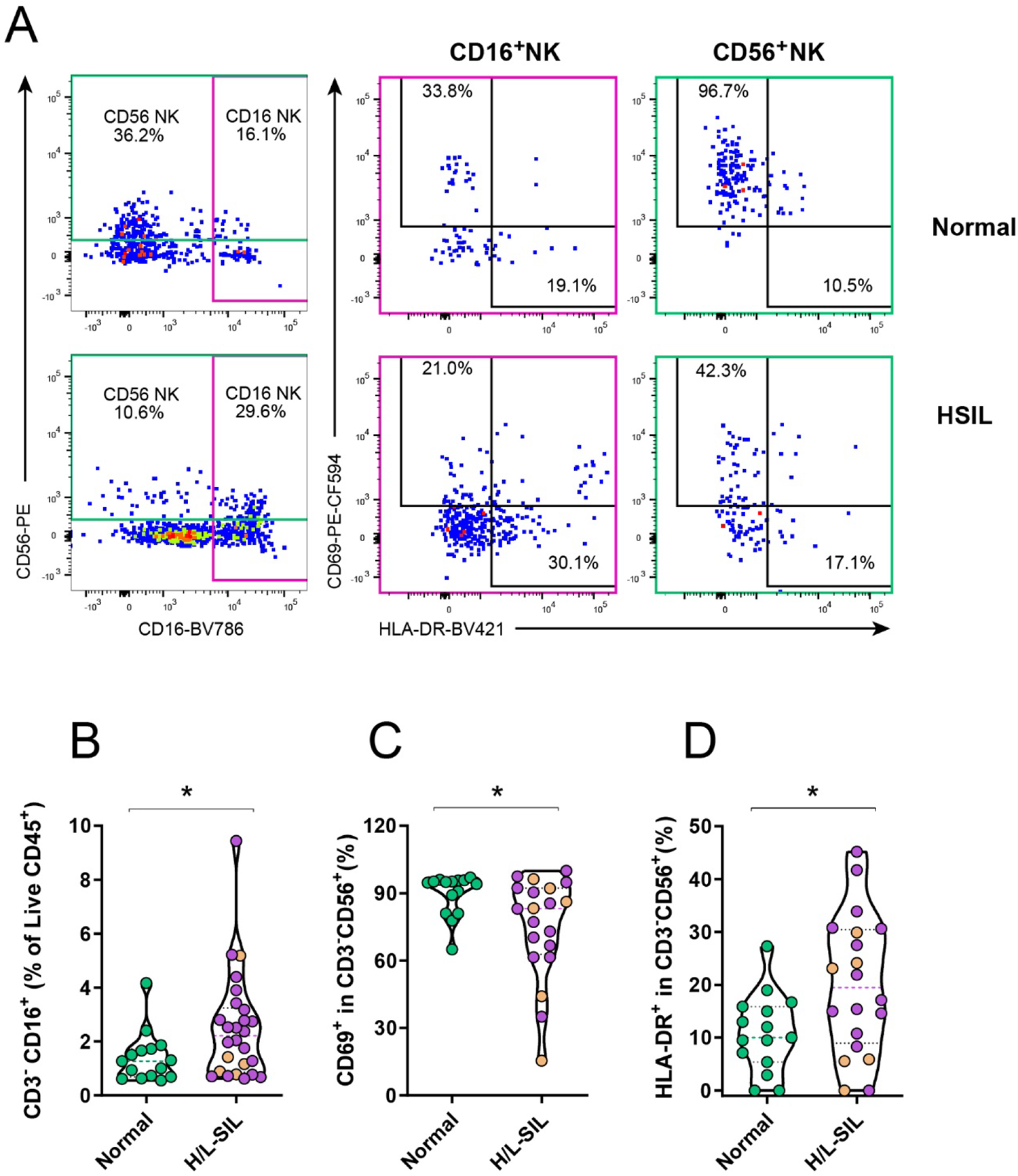
Frequency of Natural Killer lymphocyte populations associated to squamous intraepithelial lesion. **A**, Flow cytometry gating strategy used to quantify anal tissue CD3^-^ Natural Killer (NK) subsets is shown for a non-pathological sample (top) and for a high-grade squamous intraepithelial lesion sample (HSIL, bottom). Expression of CD16 (pink) and CD56 (green) was used to identify two subsets, in which the activation/resident memory marker (CD69) and the activation marker (HLA-DR) were assessed. **B**, Frequency of CD3^-^CD16^+^ NK cells out of all living CD45^+^ cells in normal (in green) *versus* pathological (H/L-SIL, in purple; HSIL are highlighted in brown) samples. **C**, Frequency of CD56^+^ NK cells expressing CD69 in normal compared to pathological tissue samples. **D**, Frequency of HLA-DR expression in CD56^+^ NK cells in normal *versus* pathological (H/L-SIL). Data are represented as a violin plot; horizontal lines are median and interquartile range. Statistical comparisons using Mann-Whitney U test for two group analyses are shown: *p<0.05.

### Potentially suppressive myeloid cell subsets augment in anal dysplasia

We additionally determined the frequency of several myeloid subsets including a subset of potentially immune tolerant cells, called myeloid-derived suppressor cells (MDSC) and mature neutrophils (CD15^+^CD16^+^; **Figure 3A**) (25, 26). Of note, MDSC were defined as CD11b^dim^ CD33^+^ myeloid cells with either an HLA-DR^-^ CD14^-^ or an HLA-DR^low^ CD14^+^ phenotype (26), as shown (**Figure 3A**). In certain samples, the percentage of MDSC expressing CD14 was significantly increased with pathology. The HSIL group showed a median of 43.85 (IQR: 34.03-54.70), and the LSIL group showed a median of 27.30 (IQR: 16.45-36.93), whereas the normal group had a median of 23.35 (IQR: 16.05-39.23). However, due to high variability within the normal samples, statistical significance was not achieved, with only a trend observed when compared to the HSIL group (p= 0.082; **Figure 3B**). Still, out of the six individuals with paired normal and LSIL samples, five of them exhibited an increase in CD14^+^MDSC in the LSIL sample compared to the non-pathological sample (**Figure 3C**). Regarding the evaluation of CD15^+^CD16^+^ neutrophils, we detected a gradual increase in their percentage associated with the severity of the SIL: there was a statistically significant difference between normal and HSIL samples (p=0.047, **Figure 3D**), and the association became stronger when all pathological samples were grouped together and compared to normal samples (p= 0.012, **Figure 3E**). Overall, increased proportions of CD14^+^MDSC and CD15^+^CD16^+^ neutrophils, both possessing potential immune suppression functions, were found to be associated with dysplasia.

**Figure 3.**
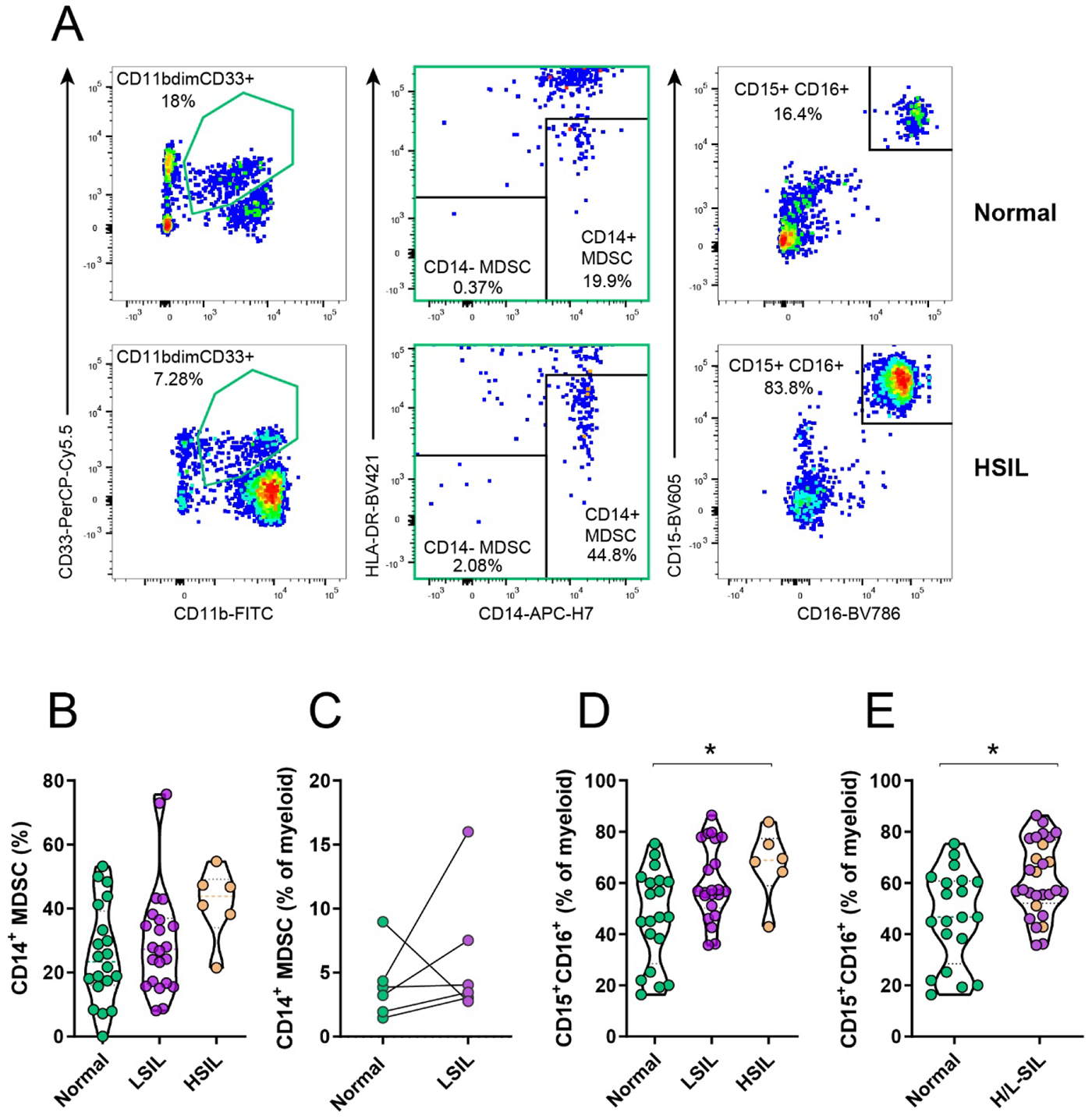
Frequency of myeloid populations associated to squamous intraepithelial lesion. **A**, Flow cytometry gating strategy used to quantify myeloid derived suppressor cells (MDSC) (out of CD11b^dim^CD33^+^, in green) and neutrophils (CD15^+^) expressing CD16 is shown for a non-pathological sample (top) and for a high-grade squamous intraepithelial lesion sample (HSIL, bottom). Sequential gating from left to right was used to determine two different MDSC subsets (CD14^-^ HLA-DR^-^ and CD14^+^ HLA-DR^dim^/^-^). **B**, Frequency of CD14^+^ MDSC out of the CD11b^dim^CD33^+^ myeloid gate in normal (green), LSIL (purple) and HSIL (brown) samples. **C**, Frequency of CD14^+^ MDSC out of total myeloid cells in paired normal and LSIL samples from the same individual analysed by the Wilcoxon signed-rank test. **D and E**, Frequency of neutrophils (CD15^+^CD16^+^) out of the total myeloid fraction in (**D**) normal, LSIL and HSIL or in (**E**) normal (in green) *versus* pathological (H/L-SIL, in purple; HSIL are highlighted in brown) samples. Data are represented as a violin plot; horizontal lines are median and interquartile range. Statistical comparisons using non-parametric Kruskal-Wallis test with Dunn’s post-hoc test for multiple comparisons and Mann-Whitney U test for two group analyses are shown: *p<0.05.

Considering that the frequency of this myeloid subset expressing CD15*^+^*CD16*^+^* appeared as the best flow cytometry-derived subset to classify pathology, we assessed the level of correlation between this parameter and other immune subsets and clinical parameters. The frequency of this subset out of the total myeloid fraction did not show any correlation with age, CD4 nadir, CD4/CD8 ratio, or the number of years under viral suppression for each individual (**Figure 4**). Nevertheless, there was a strong negative correlation between this subset and the total frequency of CD4*^+^* T_RM_ (r = – 0.41, p = 0.005) as well as CD8*^+^* T_RM_ (r = – 0.58, p < 0.005) in the same samples, the frequency of CD3*^-^* CD56*^-^* expressing CD69 (r = ȓ 0.43, p = 0.013) and the frequency of myeloid cells expressing high levels of HLA-DR (r = – 0.40, p = 0.004). In contrast, a positive correlation was observed between the total frequency of CD3*^-^* CD16*^+^* NK cells and CD15*^+^*CD16*^+^* myeloid cells (r = 0.33, p = 0.036; **Supplementary** Figure 2).

**Figure 4.**
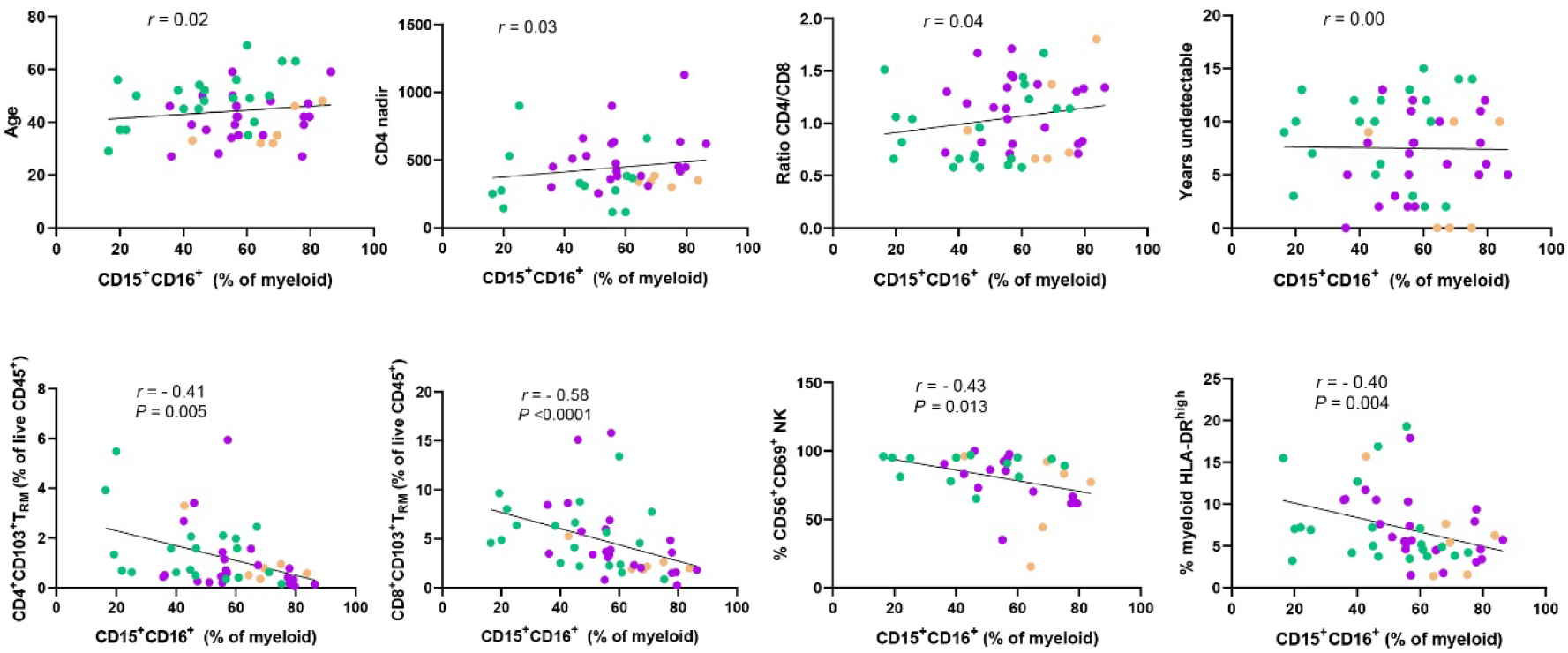
Correlation between clinical and immunological parameters and the frequency of anal CD15^+^ CD16^+^ myeloid cells in anal samples from PWH. Correlation between the frequency of CD15^+^CD16^+^ out of the total myeloid fraction in normal (green), LSIL (purple) and HSIL (brown) anal samples and clinical parameters associated to the individual (top row from left to right: age, CD4 nadir, CD4/CD8 ratio and years of being undetectable <50 copies/mL) or with other immunological subsets determined by flow cytometry and identified as defined in **Supplementary** Fig. 1, (bottom row from left to right: CD4^+^T_RM_, CD8^+^T_RM_, CD69^+^ CD3^-^ CD56^-^ NK and HLA-DR^high^ myeloid cells). Statistics were performed using non-parametric Spearman rank correlation.

### Histological confirmation of selected immune biomarkers for diagnostic

Based on our results, we selected CD103 and CD15 molecules for further confirmation of our findings through immunohistochemistry (**Figure 5A**). Our main objective was to determine their diagnostic value as individual pathology markers in comparison to p16, which is currently recommended to support the diagnosis of HSIL in the appropriate morphology context (16, 27). To this end, we obtained a new set of 57 archived tissue sections from the validation cohort (**Table 1**). CD103 and CD15-positive cells were individually counted within the epithelium or the underlying stroma. In contrast to our flow cytometry data, the average CD103 count within the epithelium and stroma of HSIL biopsies was higher compared to normal samples (**Figure 5B and C**). These findings suggested that other subsets beyond T cells, such as NK cells and CD15^+^ neutrophils, as previously reported (24, 28), could potentially exhibit a higher frequency of CD103 expression in association with pathology. Indeed, the subsequent analysis of CD103 expression within the neutrophil CD15^+^CD16^+^ subset demonstrated an overall increase of this molecule in the pathological samples compared to the normal samples (p= 0.046, **Supplementary** Figure 3).

**Figure 5.**
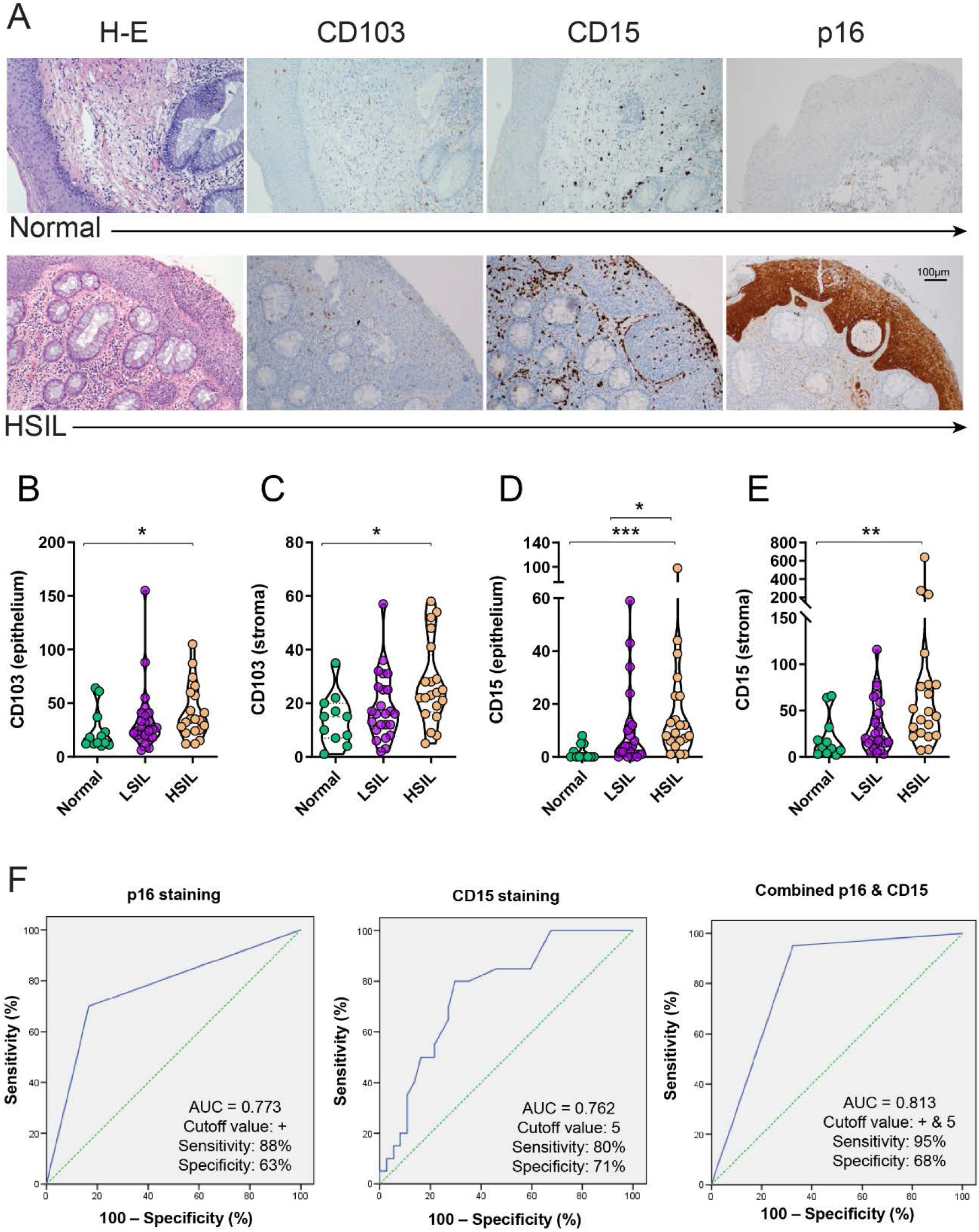
Detection of immunological biomarkers by immunohistochemistry. **A,** Examples of microphotographs showing haematoxylin and eosin, CD103, CD15 and p16 staining of the intraepithelial compartment and the underlying lamina propia (stroma) of a normal (top) and a high-grade squamous intraepithelial lesion (HSIL, bottom) mucosal samples. Original magnification 20x (scale bar is indicated). **B-E**, Quantification of the average number of positive cells detected per a median of 3 fields (range 1 to 7) at 40x of magnification in the (**B**) epithelium or the (**C**) stroma for CD103 staining, or in the (**D**) epithelium or the (**E**) stroma for CD15 staining. Data are represented as a violin plot; horizontal lines are median and interquartile range. Statistical comparisons using non-parametric Kruskal-Wallis test with Dunn’s post-hoc test for multiple comparisons are shown: *p<0.05; **p<0.01; ***p<0.001. **F,** Receiver operating characteristics curves showing the area under the curve (AUC), cut-off, sensitivity and specificity values for CD15 and p16 staining and their combination. Green dotted line: theoretical performance of an efficacy biomarker equivalent to a coin toss. Blue line: actual performance of the results

Importantly, in line with our flow cytometry immunophenotyping results, we observed an increase in CD15 counts in the epithelium and stroma of HSIL samples compared to both normal and LSIL samples (p=0.0001 and p=0.039, respectively, for the epithelium and p=0.0042 and p=0.074, respectively, for the stroma; **Figure 5D and E**). Further, in our study, p16 staining correlated with HSIL diagnosis with a sensitivity of 88% and a specificity of 63% (area under the curve, AUC 0.773, **Figure 5F**). In comparison, a threshold of > 5 positive CD15 cells in the epithelium had a sensitivity of 80% and a specificity of 71% (AUC 0.762, **Figure 5F**). Importantly, the combination of both biomarkers, meaning a threshold of > 5 positive CD15 cells or a positive p16 staining, showed a sensitivity of 95% and a specificity of 68% (AUC 0.813, **Figure 5F**). In summary, the findings from immunohistochemistry confirmed the presence of CD15 neutrophils associated with dysplasia in the anal mucosa. Moreover, the identification of these cells in the epithelium serves as a valuable pathological marker in this context.

Lastly, considering that the majority of lesions diagnosed as HSIL in the validation cohort underwent subsequent treatment, we aimed to determine the predictive value of CD15 staining regarding the response to treatment. Out of all the pathological samples that had a follow-up biopsy performed at the same previous site (19 out of the 20), three samples remained classified as HSIL, 10 samples showed a decrease to LSIL, and six samples completely responded to treatment and were classified as normal biopsies. When comparing the quantification of CD15-positive cells in the epithelium between the samples that completely responded to treatment (regressed to normality) and those that remained as an HSIL diagnosis or decreased to an LSIL, a trend towards lower numbers of this biomarker in the pathological samples that regressed was observed (**Supplementary** Figure 4). Indeed, it is noteworthy that samples negative for p16 (highlighted as triangles in **Supplementary** Figure 4) were observed in all groups with different treatment outcomes. This observation suggests that quantifying CD15 in the epithelium could potentially serve as a more reliable indicator for predicting the response to treatment, pending further validation.

## Discussion

Persistent infections share immunological features with tumor environment, where the balance between effector mechanisms and suppressive or inflammatory populations is disrupted. In this sense, the anal SIL may be of particular interest since it may combine a persistent viral infection with a tumor microenvironment. However, detailed assessment of relevant resident or infiltrated immune subsets within affected dysplastic areas has largely been missing for transitional anal tissue. Here we have studied the frequency and phenotype of several immune subsets present in this tissue to identify the immune environment associated to SIL in MSM with HIV. Overall, we identify a potentially enriched immune suppressive environment associated to pathological samples. Importantly, our findings highlight CD15 as a potential immunological marker that could contribute to an improved diagnosis of HSIL.

Effective resident immunity, including T_RM_ subsets, play essential roles in controlling persistent infections (29, 30). E6-specific CD4^+^ T cell responses may be associated with recent HSIL regression (14). In contrast, venereal wart patients, skewing of HPV-specific T cells from an effector T helper (Th) 1 to a Th2 profile or increased expression of PD-1 in infiltrated CD8^+^ T cells (31), may suggest suppressed effector immunity. Further, CD69^+^ CD103^+^ T_RM_-like cells accumulate in various human solid cancers, where they have been associated with improved disease outcome and patient survival (32). In anal dysplastic lesions, overall CD8^+^ T cell infiltration or expansion has been reported (19, 31), which concur with our observation of a higher frequency of CD8^+^ T cell lymphocytes in HSIL samples compared to simultaneous normal mucosa from the same individual. However, expression of CD103 within this compartment was lower in dysplastic compared to non-pathological samples. In this sense, a persistent depletion of CD4^+^ T cells from the mucosal compartments, including CD4^+^ T_RM_ phenotypes, has been reported in PWH who have been treated during chronic infection (11, 12). Considering that CD4^+^ T_RM_ promote the development of CD103-expressing CD8^+^T_RM_ in certain tissues (33), their generation may also be compromised in these patients. Still, in our cohort, other aspects associated to the dysplastic environment may have a greater impact, since all PWH included were ART-treated during the chronic phase. In fact, factors like transforming growth factor-beta (TGF-β) availability, which is essential for CD103 expression and T_RM_ development, epithelial dysfunction and chronic antigen exposure may affect CD103 expression (34–36). In this sense, expression of HPV oncoprotein E5 has been shown to attenuate the TGF-β1/Smad signaling, which may lead to destabilization of the epithelial homeostasis during early stages of viral infection, promoting cervical carcinogenesis (37). While we could speculate that the TGF-β signaling is affected within pathological areas (38), our data showed the opposite for mature neutrophils, which showed higher levels of CD103 expression in those areas, with more retention within the epithelium. Thus, other mechanisms such as an impaired CD38 signaling, autocrine secretion or the availability of TGF-β1 for T cells could be at play (39, 40).

NK cells are also known for their key role in viral and tumor clearance, and a decrease of circulating CD56^+^ NK cells has been associated with cervical dysplasia in HPV/HIV co-infected women (41). These cells may participate in rectal cancer remission, where higher number of CD56^+^ cells were associated with tumor regression and overall survival (42). Further, high expression of CD69 in CD56^+^ NK cells may identify resident memory NK cells in specific tissues such as the liver, lung or uterus (24, 43). Indeed, the expression of CD69 within the CD56 fraction in non-pathological samples was over >90%. However, HSIL and LSIL biopsies presented lower proportions of CD69^+^CD56^+^ NK cells, suggesting again that the shrinkage of the resident effector compartment may contribute to the lack of control of the nascent dysplasia. In contrast, HLA-DR expression and a high proportion of CD16^+^ NK cells were associated to pathology. HLA-DR indicates activation in several lymphocyte subsets, and an accumulation of HLA-DR-expressing NK cells at sites of inflammation has been reported (44). Regarding CD16^+^ NK cells, this subset includes CD56^-^CD16^+^ NK cells, which have been shown to expand during viral infections to form an anergic population with impaired cytotoxic activities (45).

Engaging effector mechanisms may be of particular importance in individuals infected with various persistent viruses, such as HPV and HIV, viruses that exploit immune modulation mechanisms from the host to induce immune tolerance and limit viral clearance (46, 47). Indeed, the local inflammatory state generated by chronic infection could induce accumulation of undesired suppressive cells, such as MDSC, as reported (48, 49). In this sense, in the progression of cervical cancer, there have been observations of increased cytokine production, including granulocyte colony-stimulating factor, which may participate in MDSC induction (50). Although previous studies suggest that myeloid cells might be displaying an immunosuppressive effect in HPV-induced malignancies (50–52), the mechanisms responsible for the various immune-related defects observed in these patients remain unclear. Furthermore, the so-called mature CD15-neutrophils, identified by high expression of CD16, may also play a controversial role. From one hand, they have been linked to inflammatory conditions, similar to what has been reported in gingival tissues of periodontitis patients (53). In this sense, neutrophil accumulation in colorectal tissue of PWH has been associated to microbiota alterations during chronic infection, which may contribute to inflammation (54). However, on the other hand, this phenotype has been shown to exert the strongest level of T cell immunosuppression (25, 50, 55). The fact that CD15^+^ granulocytic MDSC and neutrophils share expression of CD15 and CD16 molecules, indicates that only functional assays would confirm their immunosuppressive properties (56, 57), and future research in this area is warranted. Still, both CD14^+^ MDSC and CD15^+^CD16^+^ mature neutrophils are known to be key hallmarks of tumor inflammation and immune suppression, subsets that are also involved in chronic infections (25, 26). Thus, the fact that we observed a gradual increase of these subsets from normal to HSIL samples, suggests that an immune suppressive environment may favor dysplasia progression. In this sense, a link between systemic amplification of myeloid cells, and the detrimental effects of these cells on CD8^+^ T cell activation and recruitment into the tumor microenvironment has been proposed (48).

Importantly, immunohistochemistry analyses confirmed the infiltration of CD15 neutrophils in the anal mucosa associated to dysplasia, showing its potential value as a biomarker for pathology staging. Substantial disagreement exists among experienced pathologists in diagnosing SIL by H&E morphology, which is the gold standard (16). In this sense, addition of p16 immunohistochemistry increases inter-observer agreement, yet disagreement remains considerable regarding intermediate lesions (16). Thus, it is crucial to identify additional markers that can help minimize the misdiagnosis of HSIL and avoid unnecessary treatments. Moreover, the identification of reliable markers is essential for accurately identifying individuals with pre-cancerous lesions who are at risk of disease progression. In our study, the determination of CD15 and p16 showed a similar correlation with dysplasia, in addition to have a similar technical complexity, since both were detected by immunohistochemistry. Further, the combination of both markers provided a superior diagnostic value. It should also be noted that we observed an inverse association between the epithelial infiltration of CD15 in the HSIL samples and the response to treatment, which was not observed with p16. Of note, while we did not include patients with anal cancer, other works have highlighted the importance of neutrophils and CD15 expression in cancer as biomarkers of progression and response to treatment (58, 59). Thus, larger future studies should aim to validate the utility of CD15 staining as a complementary measurement for the diagnosis of L/HSIL, or even as a prognostic marker, in particular if this marker can be eventually assessed by non-invasive techniques.

It is important to note that this study is limited by the number of samples, in particular within the HSIL group in the prospective cohort, which was restricted by the complexity of the analyses and the impossibility of preselecting samples based on the degree of dysplasia. However, CD15 results were validated in the retrospective cohort, which included more homogeneous groups. Besides, because dysplasia development takes years to progress, we lack the longitudinal analyses that would inform on the predictive value of these markers regarding lesion evolution to cancer. Future studies will address the function and interactions between these resident immune cells to define key populations in anal cancer precursor progression. In summary, our results expand current knowledge of mucosal immunity in anal dysplasia. The identification of CD15 as a potential complementary biomarker for HSIL diagnosis suggests its potential application in improving diagnostic tools and may have implications for the development of targeted immunotherapeutic strategies for this condition.

## Data Availability

All data produced in the present work are contained in the manuscript.

## Acknowledgments

The authors thank the study participants. This work was primarily supported by grants from the Spanish Health Institute Carlos III (ISCIII, PI16/00736 and PI20/00160), co-funded by ERDF/ESF, “A way to make Europe”/“Investing in your future”. This work was additionally supported in part by the Spanish AIDS network Red Temática Cooperativa de Investigación en SIDA (RD16/0025/0007), the Fundació La Marató TV3 (201814-10FMTV3) and the Gilead fellowships GLD18/00008. M.J.B is supported by the Miguel Servet program funded by the Spanish Health Institute Carlos III (CP17/00179). N.M. was supported by a Ph.D. fellowship from the VHIR. The funders had no role in study design, data collection and analysis, the decision to publish, or preparation of the manuscript.

## Author Contributions

N.M., C.M., A.A-G. performed tissue processing and flow cytometry analyses. J.C. and S.L. performed the histology and immunohistochemistry analyses. J.B., A.C., J.G., V.F., collected samples and patient data. MJ.B contributed to the implementation of the study, statistical analyses and data discussion. J.B. and M.G. conceived and supervised the study, and wrote the manuscript. All authors contributed to refinement of the study protocol and approved the final manuscript.

## Notes

### Competing Interest Statement

The authors have declared no competing interest.

### Author Declarations

Informed consent for sample collection and use of information available in the medical records was obtained from all patients included. This study was performed in accordance with the Declaration of Helsinki and approved by the Institutional Review Board (PR(AG)240/2014) of the University Hospital Vall Hebron (HUVH, Barcelona, Spain).

