## Supplemental Information for "Intraepithelial CD15 infiltration identifies high grade anal dysplasia in people with HIV"

Burgos et al.

**Supplementary Table 1.** Anti-human antibodies used for flow cytometry and immunohistochemistry.

| Antibody | Flurochrome | Clone | Commercial source |
|---|---|---|---|
| **Flow Cytometry** | | | |
| CD3 | PE-Cy7 | SK7 | BD Biosciences |
| CD33 | PerCP-Cy5.5 | WM53 | BioLegend |
| CD11b | FITC | M1/70 | BioLegend |
| CD45 | Alexa700 | HI30 | BioLegend |
| CD56 | PE | B159 | BD Biosciences |
| CD14 | APC-H7 | MφP9 | BD Biosciences |
| CD8 | APC | RPA-T8 | BD Biosciences |
| CD16 | BV786 | 3G8 | BD Biosciences |
| CD103 | BV650 | Ber-ACT8 | BD Biosciences |
| CD15 | BV605 | W6D3 | BD Biosciences |
| CD20 | V500 | L27 | BD Biosciences |
| HLA-DR | BV421 | G46-6 | BD Biosciences |
| **Immunohistochemistry** | | | |
| CD15 | NA | Mouse MMA | Ventana |
| CD103 | NA | Rabbit EPR4166(2) | Abcam |
| CINtec® p16 | NA | Mouse E6H4 | Ventana |

**NA** not applicable

**Supplementary Fig. 1**

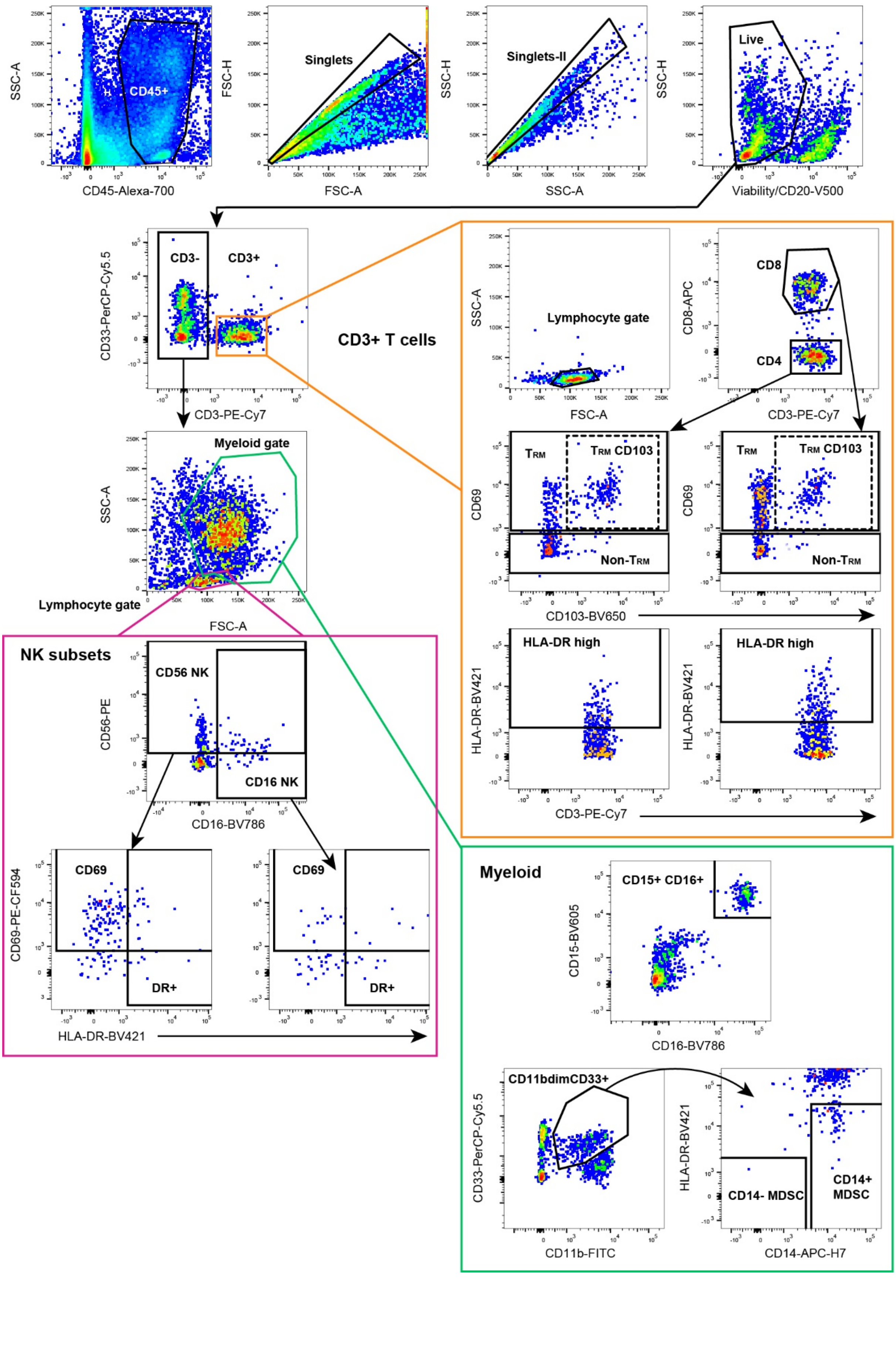

**Figure S1. Flow cytometry gating strategy used to quantify the frequencies of immunological subsets of study in anal tissue samples.** Sequential gating on the top of the figure from left to right allowed selecting single live hematopoietic non-B cells ($CD45^{+}$ $CD20^{-}$ Viability $dye^{-}$). Secondly, the expression of CD3 was used to determine $CD3^{+}$T cells ($CD3^{+}$, in orange) and natural killer (NK) or myeloid ($CD3^{-}$) lineages. For each specific lineage expression of different markers was used to determine sub-populations of interest. Within the $CD3^{+}$T cell linage, CD4 and CD8 populations were gated and their sub-populations $T_{RM}$, Non-$T_{RM}$ and $T_{RM}$ $CD103^{+}$ were determined by the use of CD69 and CD103, as well as, activated or non-activated by the expression of HLA-DR. Based on size and cellular complexity the NK (pink) and myeloid (green) linages were determined. NK lymphoid subpopulations (pink bottom left box) were defined as $CD56^{+}$ or $CD16^{+}$ and their expression of CD69 (residency and/or activation) and HLA-DR (activation) determined. Within the myeloid linage (green bottom right box), we analysed mature neutrophils determined as $CD15^{+}CD16^{+}$, as well as putative myeloid derived suppressor cells (MDSC, defined by $CD11b^{dim}CD33^{+}$), which were sequentially gated as HLA-$DR^{-}$/$CD14^{-}$ or HLA-$DR^{dim}$/$CD14^{+}$ MDSCs.

### Supplementary Fig. 2

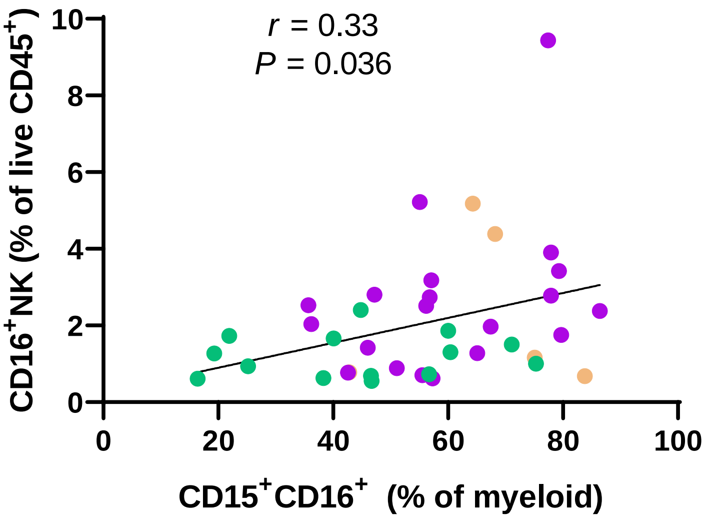


**Figure S2. Correlation between the frequency of CD16$^+$ NK cells and the frequency of anal CD15$^+$ CD16$^+$ myeloid cells in anal samples from PWH.** Correlation between the frequency of CD16$^+$ CD3$^-$ NK cells out of the total living CD45$^+$ cells and the frequency of CD15$^+$CD16$^+$ out of the total myeloid fraction in normal (green), LSIL (purple) and HSIL (brown) anal samples and the frequency if. Statistics were performed using non-parametric Spearman rank correlation and nonlinear regression.

### Supplementary Fig. 3

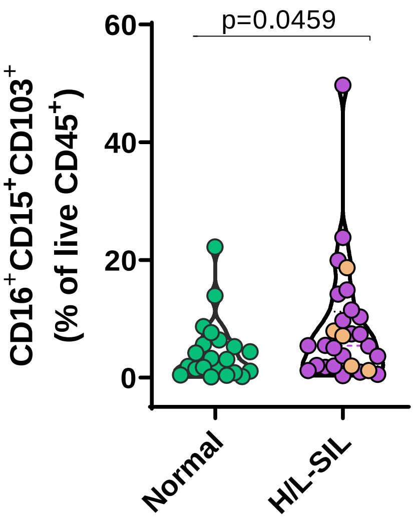


**Figure S3. Frequency of CD103$^+$ neutrophils associated to squamous intraepithelial lesion**. Frequency of neutrophils (CD15$^+$CD16$^+$) out of all living CD45$^+$ cells in normal (in green) *versus* pathological (H/L-SIL, in purple; HSIL are highlighted in brown) samples. Data are represented as a violin plot; horizontal lines are median and interquartile range. Statistical comparison using Mann-Whitney U test is shown: * $p<0.05$.

### Supplementary Fig. 4

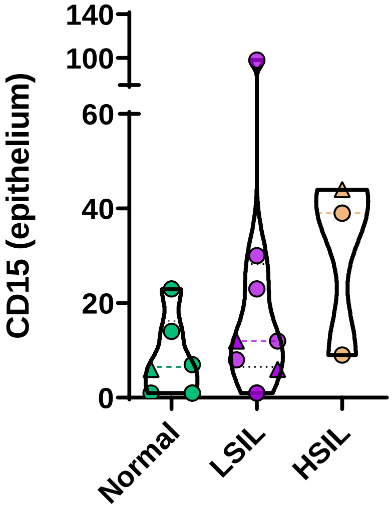


**Figure S4. Predictive value of CD15 quantification in the epithelium by immunohistochemistry.** Average number of CD15 positive cells detected per a median of 3 fields (range 1 to 7) at 40x of magnification in the epithelium of the HSIL group from the validation cohort by subsequent outcome within then same location: normal (in green), LSIL (in purple) or HSIL (in brown), in which triangles highlight samples with concomitant p16 negative staining. Data are represented as a violin plot; horizontal lines are median and interquartile range.
